# Pneumococcal vaccine uptake and vaccine effectiveness in older adults with invasive pneumococcal disease in Germany

**DOI:** 10.1101/2020.09.25.20201707

**Authors:** Stephanie Perniciaro, Mark van der Linden

## Abstract

**Background:** Invasive pneumococcal disease (IPD) in people ≥60 years old is on the rise in Germany. There has been a recommendation for pneumococcal vaccination in this age group since 1998.

**Methods:** We determined the vaccination status of people ≥60 years old with IPD in Germany. We assessed vaccine effectiveness (VE) of the recommended 23-valent polysaccharide vaccine (PPV23).

**Results:** The rate of pneumococcal vaccination in older adults with IPD is low, 26%, with only 16% of people receiving a pneumococcal vaccine within the past 5 years. Age- and sex-adjusted vaccine effectiveness (VE) for PPV23 was 37% (95% confidence interval 12% - 55%). For people vaccinated with PPV23 less than 2 years prior to IPD, VE was - 20% (−131% - 34%), between 2-4 years prior to IPD, VE was 56% (20% - 76%), and 47% (17% - 63%) for those vaccinated ≥5 years ago. Excluding serotype 3, overall VE for the remaining serotypes in PPV23 was 63% (49% - 74%). For people receiving PPV23 within the past 2 years, VE against all serotypes except 3 was 49% (12% - 71%); for people vaccinated between 2-4 years prior to IPD 66% (37% - 82%); for those vaccinated ≥5 years ago, 69% (50% - 81%). VE of PPV23 against serotype 3 IPD only was -110% (- 198% - -47%).

**Conclusions:** To reduce IPD in older adults in Germany, we must increase the rate of pneumococcal vaccine uptake. For 22/23 serotypes, PPV23 was effective. Serotype 3 remains a major problem.

## Introduction

The worldwide disease burden of invasive pneumococcal disease among the elderly remains high. The most common presentation of pneumococcal disease in this age group is community acquired pneumonia (CAP). And although the larger part of CAP is non-bacteremic, an estimated 3-6% ^1,2^ of CAP cases progress to invasive disease, with pneumococci entering the blood stream (bacteremic pneumonia). An estimated 50% of pneumonia in the elderly is caused by pneumococci,^3–5^ with serotype 3 as the most prevalent serotype.^6–8^ Pneumococcal polysaccharide vaccines were introduced as early as 1977 (a 14-valent vaccine, PPV14) and 1983 (a 23-valent vaccine, PPV23),^9,10^ but incidence and mortality of pneumococcal disease in older adults have remained largely unchanged. Apart from the broadly discussed effectiveness of PPV23, which is currently recommended for the elderly in many countries, there are reports of low uptake of adult pneumococcal vaccination.^11,12^

To address this important cause of morbidity and mortality in older adults in Germany, the National Immunization Technical Advisory Group (STIKO) of the Robert Koch Institute first recommended that all adults 60 years and older receive a dose of PPV23 in 1998. The recommendation has been reassessed periodically, and currently calls for vaccination with PPV23 for all adults 60 years and older, with possible repeat doses at ≥6-year intervals according to perceived necessity (Supplemental Table 1). Individuals with congenital immune defects or immune suppression, as well as individuals with increased risk for pneumococcal meningitis are recommended to receive sequential vaccination with PCV13 followed by PPV23 6-12 months later. Individuals with other chronic diseases are recommended one dose of PPV23. ^13^

There is not a single-product national immunization plan for pneumococcal vaccines in Germany: individual providers (in consultation with their patients) are allowed to choose which vaccine is used (within the recommendations) after which reimbursement by health insurance companies is guaranteed. STIKO issues general recommendations for all German federal states. However, some federal states also have their own vaccine committees, which can issue different recommendations. One federal state out of 16 (Saxony), representing 5% of the German population, has recommended sequential vaccination with the 13-valent pneumococcal conjugate vaccine (PCV13) followed by PPV23 for all adults 60 years and older with and without underlying health conditions, though PCV13 costs are not reimbursed. ^14^

Considering the forthcoming higher-valent pneumococcal conjugate vaccines ^15,16^ and the increasing burden of IPD in this age group, ^17^ the recommendation may be re-evaluated soon. In this study, we wished to determine the rate of pneumococcal vaccine uptake among older adults with IPD, and assess vaccine effectiveness (VE) in this population.

## Materials and Methods

### Surveillance Methods

We performed a two-year prospective survey of pneumococcal and influenza vaccination status for adults ≥ 60 years old with IPD that sent a pneumococcal isolate to the German National Reference Center for Streptococci (GNRCS) between January 1, 2018 and December 31, 2019. The GNRCS surveillance system is estimated to receive around half of all invasive pneumococcal isolates in Germany. ^18^ Germany lacks a central patient registry, so for all pneumococcal isolates originating from IPD cases in people 60 years and older, we called clinical laboratories, hospital admissions departments, and primary care providers to ascertain the vaccination status of patients with IPD.

### Analytical Methods

Pneumococcal isolates were identified and serotyped as previously described {van der Linden, 2019 #5181} using Neufeld’s Quellung method with sera from the Statens Serum Institute.

Records of IPD cases were grouped by vaccination status (unknown, not vaccinated, vaccinated with PCV13, vaccinated with PPV23, vaccinated with both PCV13 and PPV23). VE was calculated by comparing the vaccination status of people with vaccine-serotype (VT) IPD to the vaccination status of people with non-vaccine serotype (NVT) IPD. ^19^ VE was adjusted for age and sex with Firth’s bias-reduced logistic regression ^20,21^ where VT IPD was the dependent variable and the patient’s vaccination status, age in years, and sex were independent variables.

We also separated groups which had been vaccinated less than 2 years prior to onset of IPD, between 2-4 years prior to IPD, and those vaccinated ≥5 years prior to IPD and repeated the analyses. We also grouped people by age, from 60-69, 70-79, and ≥80, and repeated the analyses. VT IPD was defined for PPV23 as serotypes (1, 2, 3, 4, 5, 6B, 7F, 8, 9N, 9V, 10A, 11A, 12F, 14, 15B, 17F, 18C, 19F, 19A, 20, 22F, 23F, and 33F) and for PCV13 as serotypes (1, 3, 4, 5, 6A, 6B, 7F, 9V, 14, 18C, 19A, 19F, and 23F). Because of previous work ^3,22,23^ indicating poor VE against serotype 3, we also calculated VE for all serotypes except 3, and for serotype 3 separately. We assessed correlation between pneumococcal vaccination status and influenza vaccination status with Spearman correlations. All analyses were performed in R (The R Foundation for Statistical Computing, Version 3.6.1) using the package *logistf*. ^21^

## Results

From January 1, 2018 to December 31, 2019 the GNRCS received 6310 IPD isolates from individuals 16 years and older. Of these, 4642 (74%) were samples from IPD in people 60 years and older. We were able to determine the vaccination status for 928 people (20%), 243 (26%) of whom had ever received a pneumococcal vaccine and 146 (16%) of whom had received a pneumococcal vaccine within the past five years (Figure 1). 67 people who were not vaccinated at the time of their IPD episode were then vaccinated after recovery.

**Figure 1.**
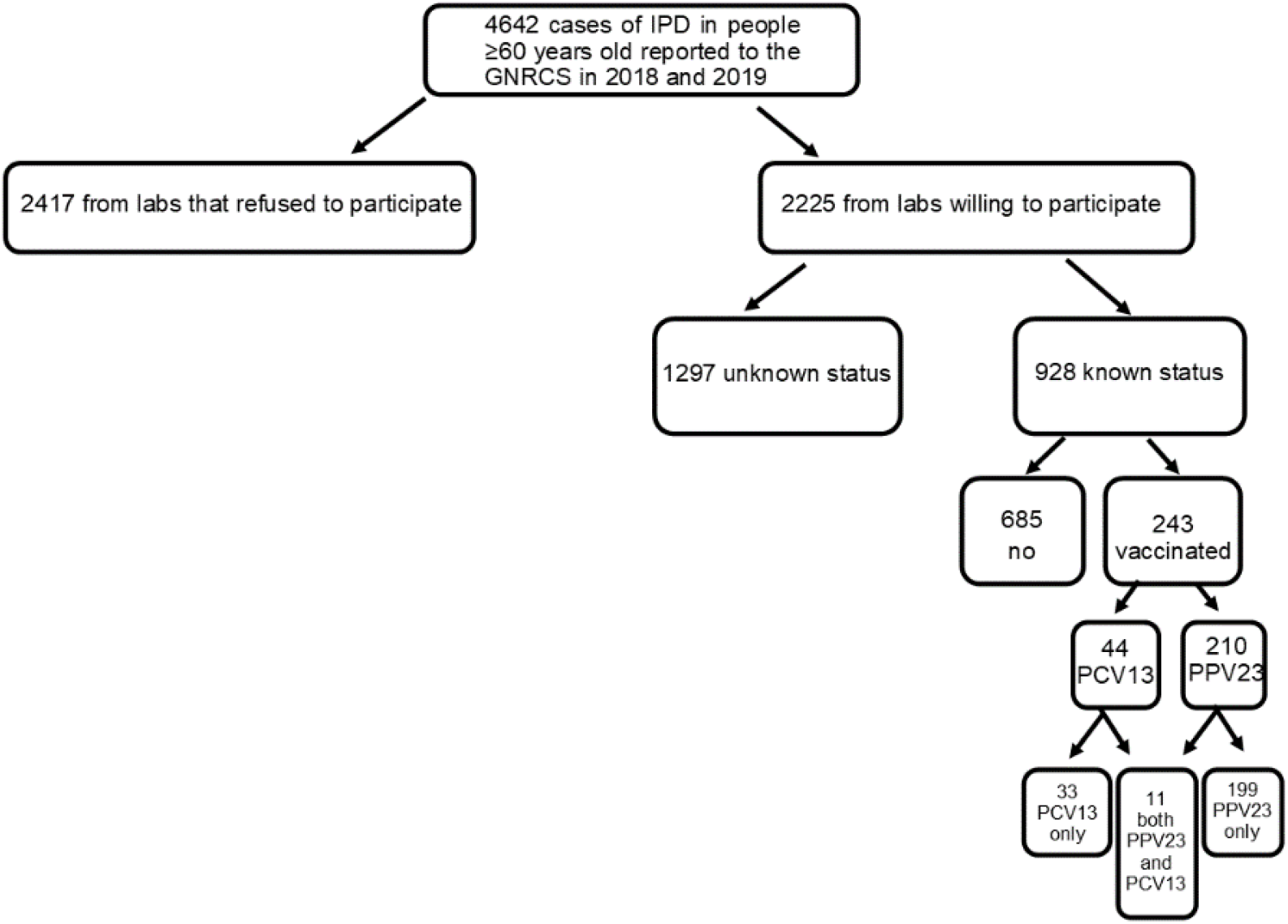
Study population of invasive pneumococcal disease cases from people ≥60 years old in Germany, 2018-2019. 928 cases were used in the full analysis for vaccine effectiveness calculations.

We compared the serotype distribution of isolates for which we could not determine a vaccination status with isolates with a known vaccination status. There appeared to be no major differences in the serotype prevalence (Figure 2).

**Figure 2.**
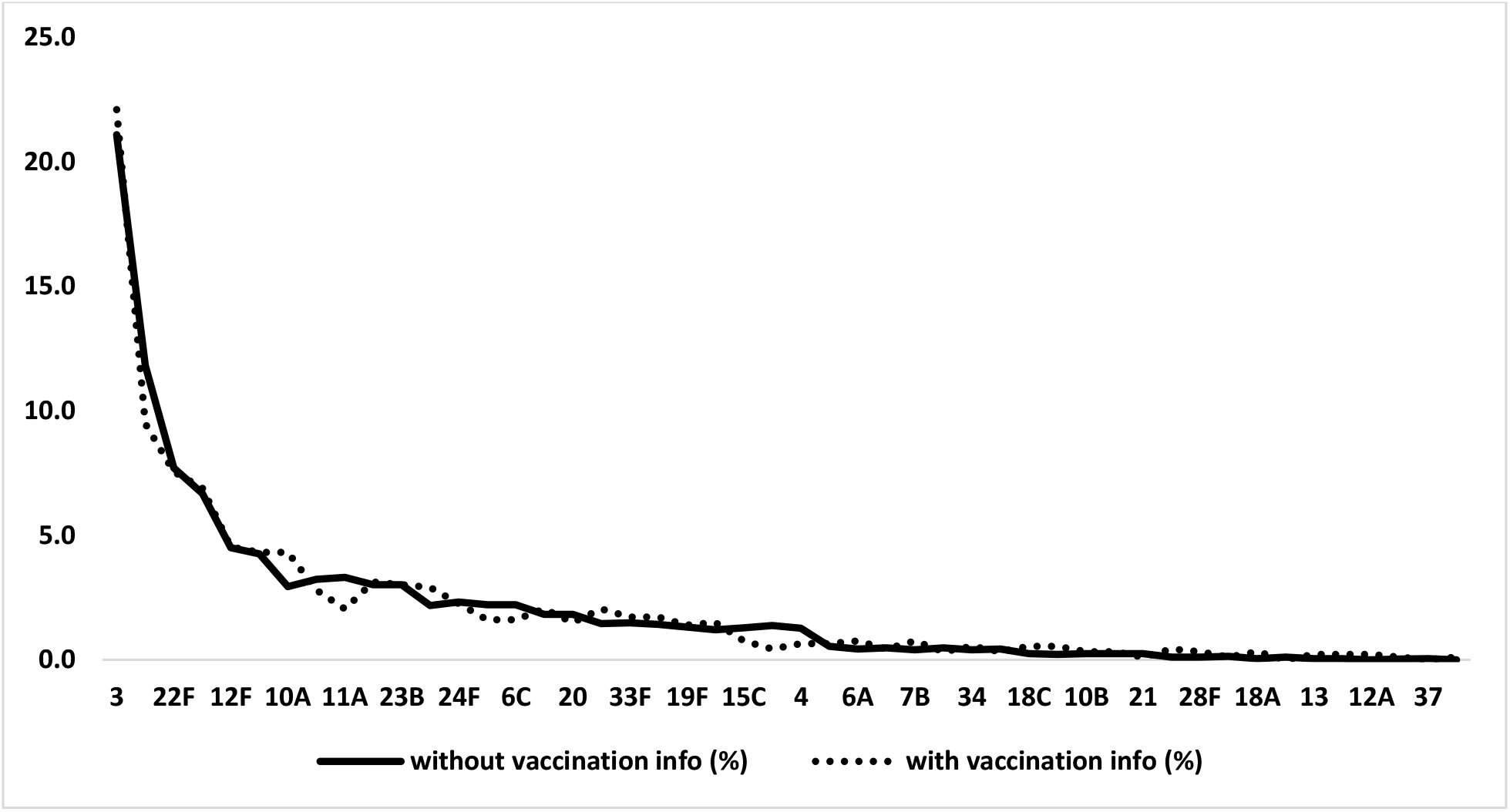
Percentage of serotypes in the GNRCS collection from adults ≥60 with IPD, by study participation. The black line represents the serotype distribution of IPD cases for which we were not able to determine a vaccination status; the light gray line represents cases for which we were able to determine a vaccination status.

We calculated age- and sex-adjusted VE for VT IPD for all PPV23 serotypes, for serotype 3 individually, and for PPV23 serotypes excluding serotype 3 (Table 1) and repeated the process for PCV13 (Supplemental Table 2). PPV23 had an overall VE of 37% (95% confidence interval (CI) 12% - 55%, a VE of -110% (CI: -198% - -47%) against serotype 3 IPD, and a VE of 63% (CI: 49% - 74%) against all PPV23 serotypes except 3. Calculations of PPV23 VE against individual serotypes were limited by small sample sizes (Supplemental Table 3), but VE against serotype 12F IPD was 87% (CI: 52-99%) and 46% (CI: 1-73%) against serotype 8 IPD.

**Table 1:** Age- and sex-adjusted vaccine effectiveness (VE) of PPV23 against IPD from PPV23 serotypes, PPV23 serotypes except 3, serotype 3, and PPV23 serotypes not included in PCV13 overall, and divided into groups based on time since vaccination: vaccinated <2 years ago, between 2-4 years ago, or ≥5 years ago. Age and sex were adjusted using Firth’s bias-reduced logistic regression; VE was calculated by the indirect cohort method. ^19^

|  | VE (%) | LL(%) | UL(%) |
| --- | --- | --- | --- |
| PPV23 serotypes | 37 | 12 | 55 |
| PPV23 serotypes except serotype 3 | 63 | 49 | 74 |
| serotype 3 | -110 | -198 | -47 |
| PPV23-nonPCV13 serotypes | 59 | 42 | 72 |
| PPV23 serotypes (vaccinated <2 years ago) | -20 | -128 | 34 |
| PPV23 serotypes (vaccinated between 2 and 4 years ago) | 56 | 20 | 76 |
| PPV23 serotypes (vaccinated ≥5 years ago) | 47 | 17 | 66 |
| PPV23 serotypes except serotype 3 (vaccinated <2 years ago) | 49 | 12 | 71 |
| PPV23 serotypes except serotype 3(vaccinated between 2 and 4 years ago) | 66 | 37 | 82 |
| PPV23 serotypes except serotype 3 (vaccinated ≥5 years ago) | 69 | 50 | 81 |
| PPV23 serotypes, Age 60-69 | 71 | 42 | 86 |
| PPV23 serotypes except serotype 3, Age 60-69 | 67 | 33 | 85 |
| Serotype 3, Age 60-69 | -1 | -117 | 56 |
| PPV23 serotypes, Age 70-79 | 28 | -27 | 58 |
| PPV23 serotypes except serotype 3, Age 70-79 | 63 | 38 | 79 |
| Serotype 3, Age 70-79 | -159 | -361 | -45 |
| PPV23 serotypes, Age ≥80 | 12 | -53 | 49 |
| PPV23 serotypes except serotype 3, Age ≥80 | 60 | 32 | 77 |
| Serotype 3, Age ≥80 | -176 | -390 | -55 |

Separating PPV23-vaccinated people by time since vaccination revealed that 30% of people ≥60 with IPD had been vaccinated within the last two years, and separating PPV23-vaccinated people by age group showed that 23% of people 60-69 with IPD had been vaccinated ≥6 years ago, 45% of people 70-79 had been vaccinated ≥6 years ago, and 44% people 80 and over had not been vaccinated within the last 6 years. Serotype distribution by vaccination status (Figure 3) and for PPV23-vaccinated people by age group (Figure 4) show that serotype 3 remains a dominant cause of IPD for both vaccinated and unvaccinated people ≥60 and older.

**Figure 3.**
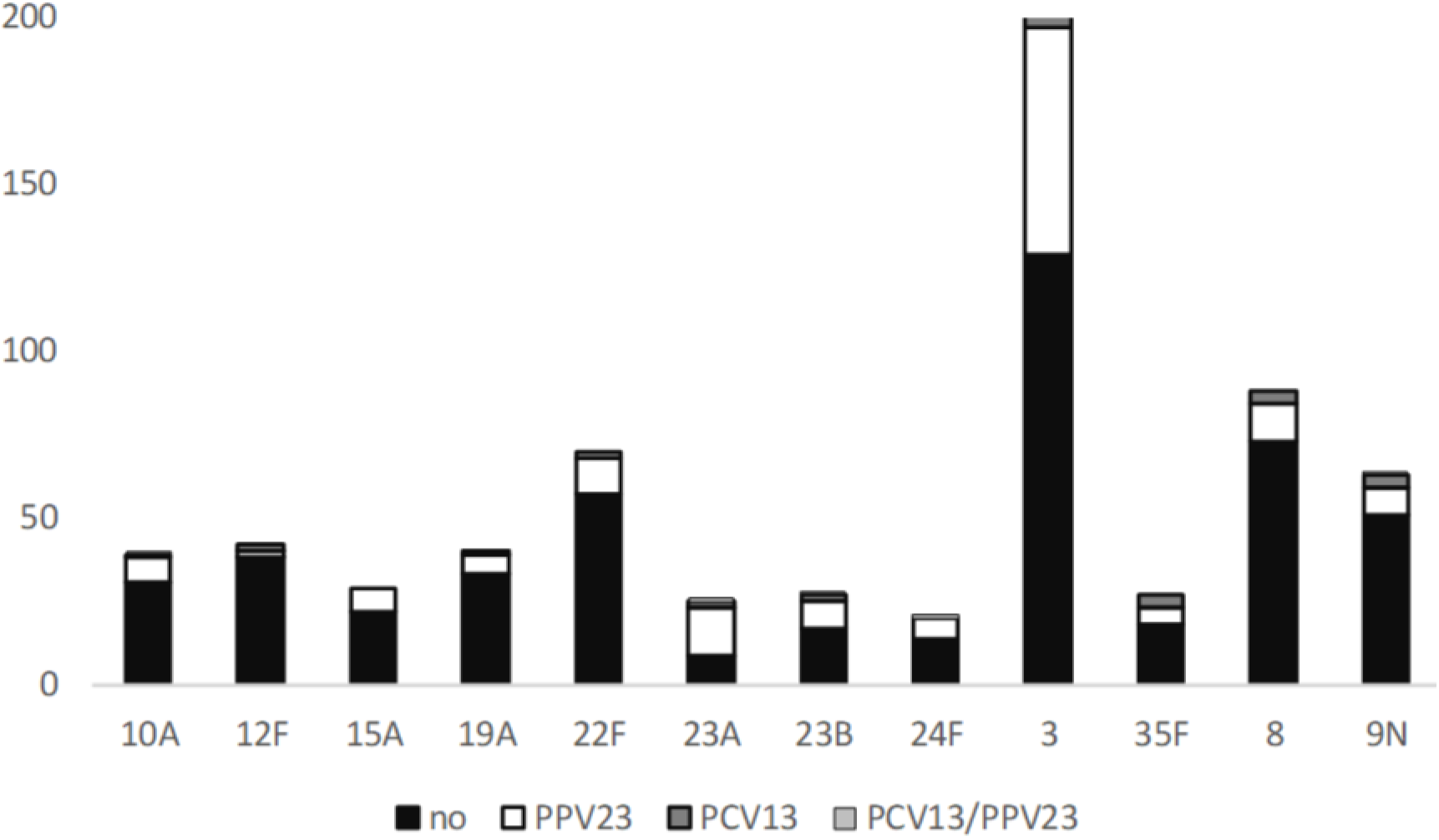
Invasive pneumococcal disease serotype distribution in adults ≥60 by vaccination status, either unvaccinated “no”, vaccinated only with PPV23, “PPV23”, vaccinated only with PCV13, “PCV13”, or vaccinated with both PPV23 and PCV13 “PCV13/PPV23”. The twelve most frequently occurring serotypes are shown.

**Figure 4.**
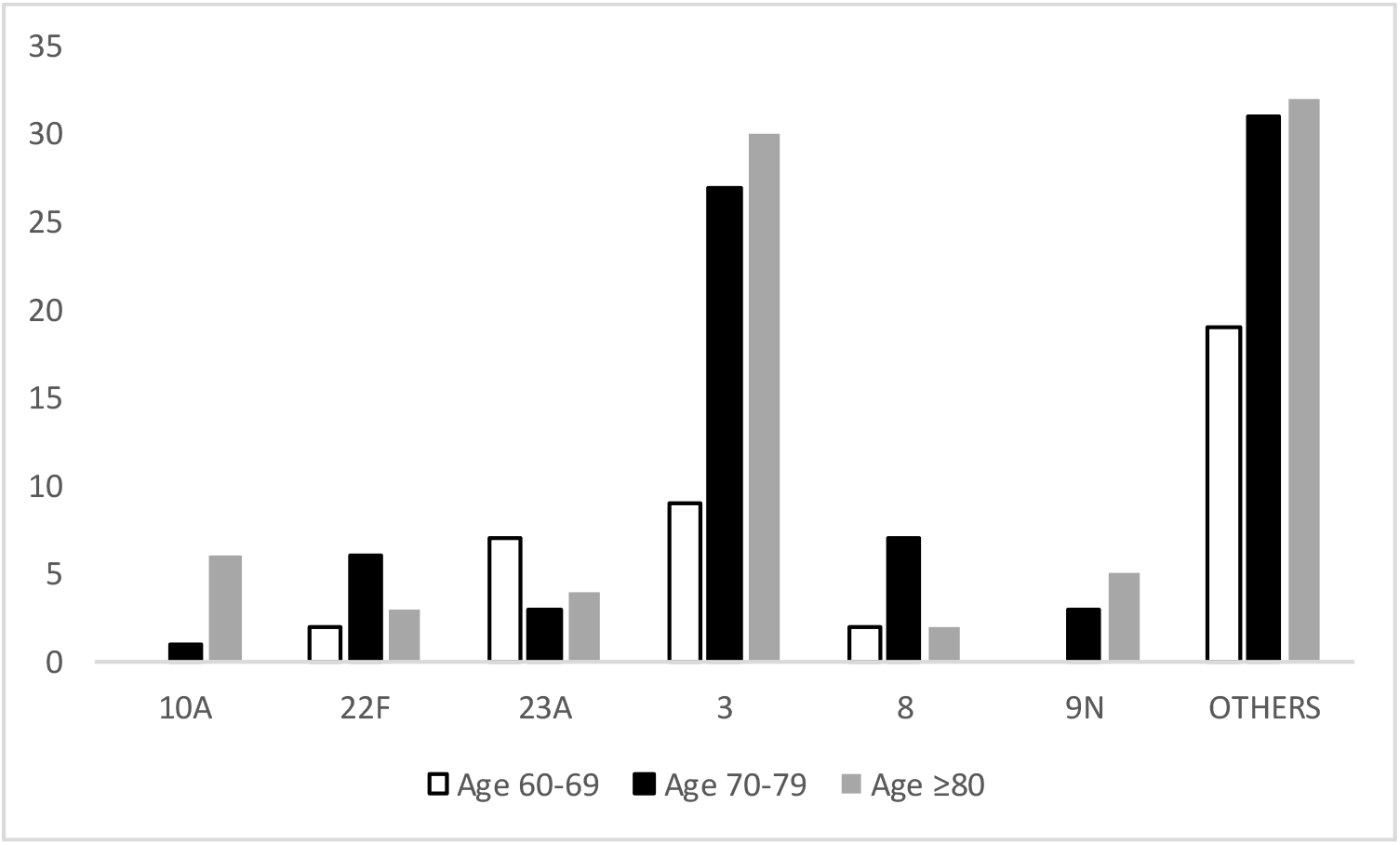
Serotype distribution by age group in people with IPD who were vaccinated with PPV23.

Vaccination with either PPV23 or PCV13 correlated strongly with receiving an influenza vaccination (Figure 5).

**Figure 5.**
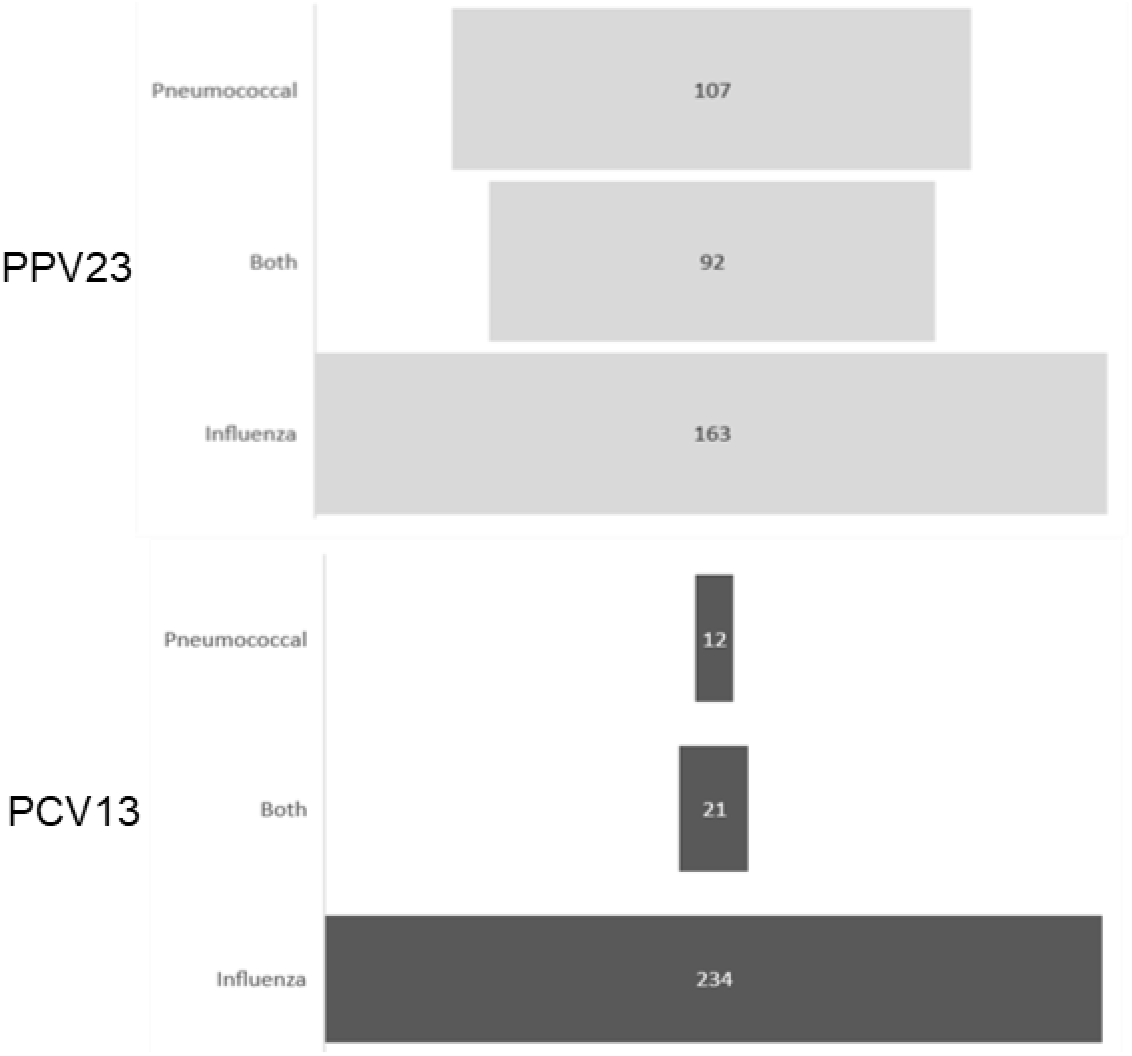
Concurrence between pneumococcal and influenza vaccination in adults ≥60 with IPD in Germany. PPV23 is shown in the top graph (light gray); PCV13 is shown on the bottom graph (dark gray).

## Discussion

### Rate of pneumococcal vaccination

Recent estimates of adult vaccination in Germany indicate that pneumococcal vaccination rates are low, and our study falls in line with these expectations. Rates of pneumococcal vaccination in the general population of adults aged ≥60 were between 4.2% and 28.4% in 2015-2019. Our study found that 26% of people with IPD aged ≥ 60 had received a pneumococcal vaccine, which indicates that our study population had similar vaccination behavior to the general population of older adults in Germany. ^24,25^ Even among vaccinated people, adherence to schedule recommendations were poor: nearly half of people ≥70 years old had not been vaccinated within 6 years of their IPD episode, and over a third of people had not been vaccinated within the past decade.

Work is underway to develop effective methods of communicating with this age group and their primary care providers about vaccination. ^26^ A recent survey of German family practice physicians indicated that physicians’ own vaccination behaviors influenced their likelihood of recommending vaccines to their adult patients, and their likelihood of remembering to check patients’ vaccination status. Encouraging or incentivizing physicians to get vaccinated themselves may be an avenue to improve the low adult vaccination rates in Germany. ^27^ The strong correlation found between pneumococcal vaccination and influenza vaccination may provide an opportunity to increase pneumococcal vaccination rates, and the potential of bundling influenza and pneumococcal vaccines should be further investigated. ^26^ Before we can realistically expect further population-level reductions in VT IPD in older adults, we must engage with this age group and their care providers to increase the rate of vaccination.

### Vaccine effectiveness

The overall VE of PPV23 established in this study was substantially higher than that found in the UK ^22^ and South Korea, ^28^ but similar to VE described in Japan, ^29^ and lower than the results in two meta-analyses. ^9,30^ The VE of PPV23 excluding serotype 3 was high (63%), and in concordance with values published earlier. Our study corroborates the total lack of protection of PPV23 against serotype 3. ^22^ PPV23 even has a strongly negative VE, possibly indicating an increased risk for vaccinated individuals to serotype 3 IPD. This finding is of particular importance, as serotype 3 VE is a main driver in the cost-effectiveness model for the German vaccine recommendation. ^9^

VE of PCV13 against serotype 3 IPD is variable, with wide-ranging estimates appearing in different populations and study methods. ^3,23^ In our study, PCV13 was only used in 33 patients with IPD (3.5%), and low VT sample sizes resulted in unreliable estimates with very wide confidence intervals, so we are unable to make definitive conclusions about PCV13 VE in the older adult population in Germany.

### Policy considerations

Combined with the reported lack of herd protection of the childhood PCV13 vaccination on serotype 3 IPD among adults in Germany, and the fact that currently 20% of IPD in adults 60 years and older in Germany is caused by serotype 3, ^17^ the negative VE found for PPV23 against serotype 3 IPD indicates that the current pneumococcal vaccination strategy for older adults does not offer sufficient protection against one of the major causes of both IPD and non-bacteremic pneumonia in this age group. The primary problem with the current pneumococcal vaccination recommendation is low uptake, but if increasing uptake does not address the burden of disease caused by serotype 3, the current recommendation will not adequately address the needs of this growing population group in Germany.

Around half of the isolates received in this study were sent by clinical laboratories which refused to participate in the study, many of them citing a recently enacted general data protection regulation (GDPR) issued by the European Union Despite the law having specific provisions for the sharing of sensitive information for public health surveillance purposes, ^31^ there is evidently some confusion about the interpretation and implementation of these new requirements. Additional official guidance is needed to inform clinical laboratories how to safely provide data in order to continue infectious disease surveillance activities so that valuable data are not needlessly lost.

## Conclusions

PPV23 offers moderate protection overall, but solid protection against PPV23 serotypes except serotype 3. PCV13 is underutilized in adults in Germany. In order to accurately assess pneumococcal vaccine impact in older adults in Germany, pneumococcal vaccine uptake must be increased.

## Data Availability

Full data for this study will be made available on request.

## Acknowledgements

We thank all of the participating laboratories, hospitals and primary care providers, the Center for Translational & Clinical Research (CTC-A) at the University Hospital RWTH Aachen, and Jutta Krasenbrink.

## Supplemental Materials

**Supplemental Table 1.**
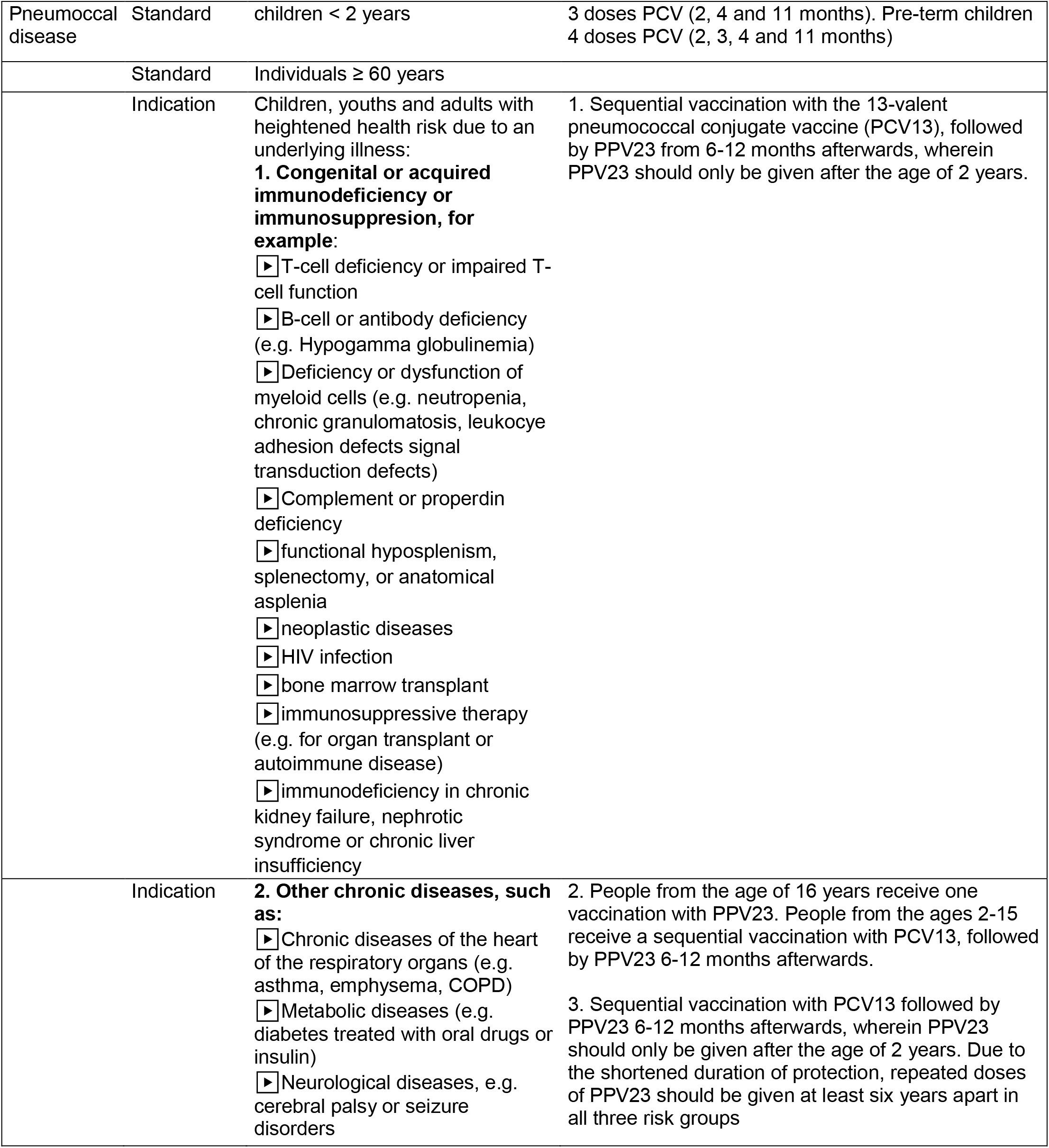
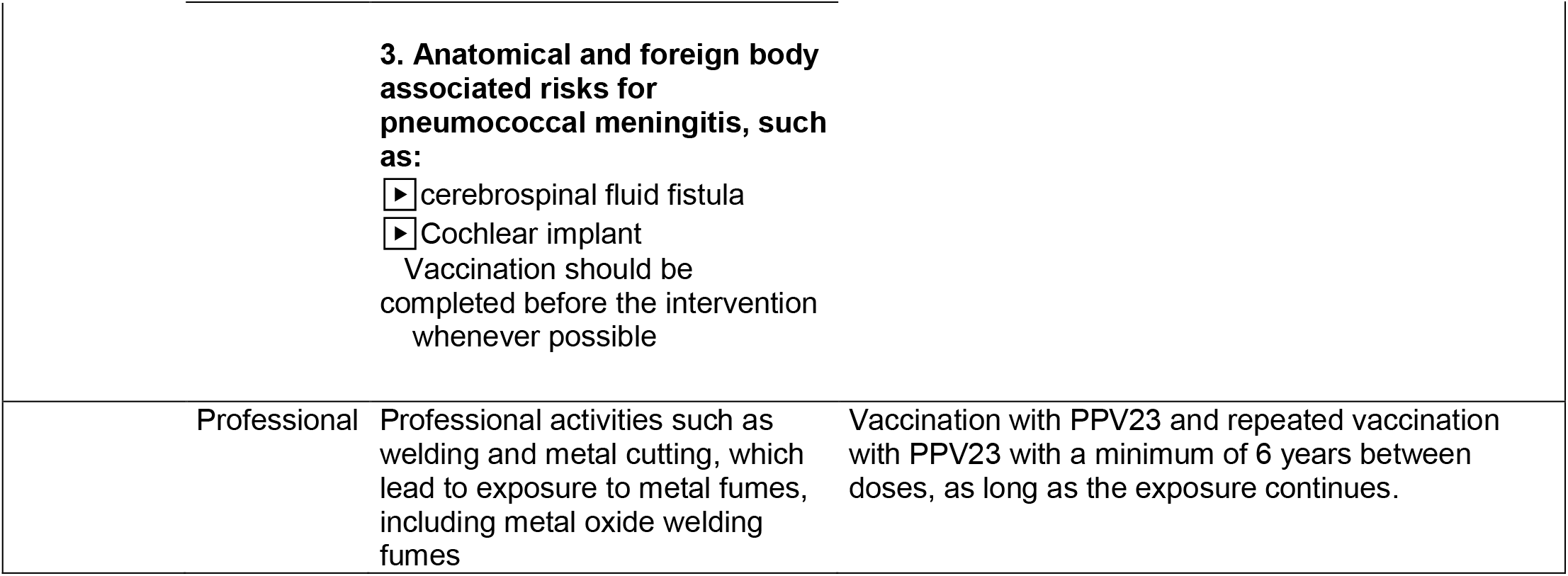
German pneumococcal vaccination recommendations by risk category

**Supplemental Table 2.**
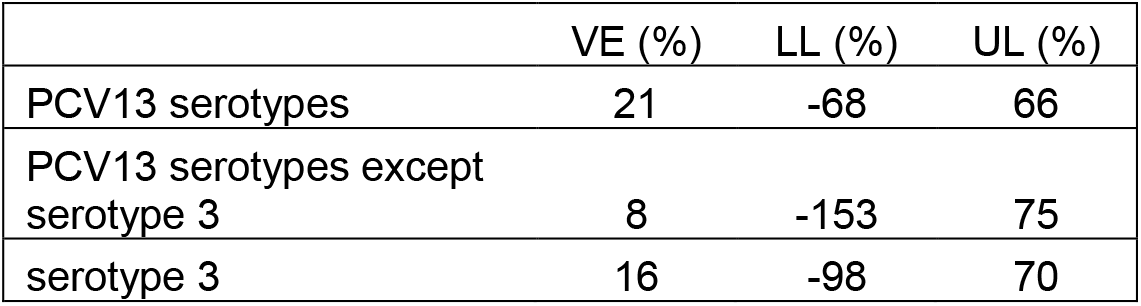
Age– and sex– adjusted vaccine effectiveness (VE) of PCV13 IPD. Lower and upper bounds of 95% confidence intervals are shown (LL, UL)

|  | VE (%) | LL (%) | UL (%) |
| --- | --- | --- | --- |
| PCV13 serotypes | 21 | -68 | 66 |
| PCV13 serotypes except serotype 3 | 8 | -153 | 75 |
| serotype 3 | 16 | -98 | 70 |

**Supplemental Table 3.** Age– and sex– adjusted vaccine effectiveness (VE) of PPV23 for single serotype IPD. Lower and upper bounds of 95% confidence intervals are shown (LL, UL)

|  | VE (%) | LL (%) | UL(%) |
| --- | --- | --- | --- |
| 10A | 20 | -73 | 67 |
| 12F | 87 | 52 | 99 |
| 19A | 36 | -42 | 75 |
| 22F | 34 | -23 | 67 |
| 3 | -110 | -198 | -47 |
| 8 | 46 | 1 | 73 |

